# Elucidating the genetic architecture of DNA methylation to identify promising molecular mechanisms of disease

**DOI:** 10.1101/2022.04.13.22273848

**Authors:** Jiantao Ma, Roby Joehanes, Chunyu Liu, Amena Keshawarz, Hwang Shih-Jen, Helena Bui, Brandon Tejada, Meera Sooda, Peter J. Munson, Demirkale Y. Cumhur, Paul Courchesne, Nancy L. Heard-Costa, Achilleas N. Pitsillides, Mike Feolo, Nataliya Sharopova, Ramachandran S. Vasan, Tianxiao Huan, Daniel Levy

**Affiliations:** Division of Nutrition Epidemiology and Data Science, Friedman School of Nutrition Science and Policy, Tufts University, Boston, MA; Population Sciences Branch, National Heart, Lung, and Blood Institute, National Institutes of Health, Bethesda, Maryland and the Framingham Heart Study, Framingham, MA; Department of Biostatistics, School of Public Health, Boston University, Boston, MA; Boston University and National Heart, Lung, and Blood Institute Framingham Heart Study, Framingham, MA; Critical Care Medicine Department, Clinical Center, National Institutes of Health, Bethesda, MD; Boston University School of Medicine, Boston University, Boston, MA; National Center for Biotechnology Information, Bethesda, MD; Department of Ophthalmology and Visual Sciences, University of Massachusetts Medical School, Worcester, MA

## Abstract

DNA methylation commonly occurs at cytosine-phosphate-guanine sites (CpGs) that can serve as biomarkers for many diseases. We analyzed whole genome sequencing data to identify DNA methylation quantitative trait loci (mQTLs) in 4,126 Framingham Heart Study participants. Our mQTL mapping identified 94,362,817 *cis*-mQTLvariant-CpG pairs (for 210,156 unique autosomal CpGs) at *P*<1e-7 and 33,572,145 *trans*-mQTL variant-CpG pairs (for 213,606 unique autosomal CpGs) at *P*<1e-14. Using *cis*-mQTL variants for 1,258 CpGs associated with seven cardiovascular disease risk factors, we found 104 unique CpGs that colocalized with at least one cardiovascular disease trait. For example, cg11554650 (*PPP1R18*) colocalized with type 2 diabetes, driven by a single nucleotide polymorphism (rs2516396). We performed Mendelian randomization (MR) analysis and demonstrated 58 putatively causal relations of CVD risk factor-associated CpGs to one or more risk factors (e.g., cg05337441 [*APOB*] with LDL; MR *P*=1.2e-99, and 17 causal associations with coronary artery disease (e.g. cg08129017 [*SREBF1*] with coronary artery disease; MR *P*=5e-13). We also showed that three CpGs, e.g., cg14893161 (*PM20D1*), are putatively causally associated with COVID-19 severity. To assist in future analyses of the role of DNA methylation in disease pathogenesis, we have posted a comprehensive summary data set in the National Heart, Lung, and Blood Institute’s BioData Catalyst.

## Introduction

DNA methylation, the most frequently studied epigenetic modification, involves the transfer of a methyl group to the fifth carbon position of the cytosine DNA nucleotide to form 5-methylcytosine.^1^ DNA methylation is influenced both by genetic and environmental factors and may mediate gene-environment interactions; therefore, it may be used to determine the risk of many complex diseases through its critical role in gene expression regulation.^2,3^ Associations between DNA methylation and a wide range of phenotypes have been identified by epigenome-wide association studies (EWAS).^4-6^ DNA methylation therefore can serve both as biomarkers for disease and contribute to its pathogenesis.

Identification of genetic loci associated with the methylation of cytosine-phosphate-guanine sites (CpGs) – i.e., DNA methylation quantitative trait loci (mQTLs) – can facilitate the interpretation of the biological underpinnings regarding the DNA methylation and disease relations and causal inference regarding the roles of DNA methylation in disease. Genome-wide association studies (GWAS) have successfully identified many disease-associated genetic variants.^7^ Molecular mechanisms linking these variants to disease, however, are not fully understood. Exploring colocalization of disease-associated genetic variants from GWAS with mQTL variants may further reveal molecular mechanisms underlying the associations between genetic variants and diseases.^8^ We hypothesize that by studying the overlap of mQTL variants with known disease-associated genetic variants from GWAS, we can further explore the joint contributions of genetic and environmental influences to diseases. Furthermore, by colocalizing mQTLs with genetic variants associated with gene expression (expression quantitative trait loci, eQTLs), we can better interpret the biological functions of disease-associated CpGs.^8^ Utilizing effect sizes derived from GWAS for mQTL variants with different diseases, we can conduct causal inference testing to explore the putative causal roles of CpGs on a wide range of diseases.^9-14^

In our earlier work, we performed GWAS of ∼415,000 CpGs in whole blood derived DNA in Framingham Heart Study (FHS) participants with validation in the Atherosclerosis Risk in Communities (ARIC) study and the Grady Trauma Project (GTP).^15^ Genotyping was performed using commercial arrays with imputation across the genome. The present study greatly expands on our prior work by incorporating whole genome sequencing (WGS) data in FHS participants obtained as part of the National Heart, Lung, and Blood Institute’s (NHLBI) Trans-Omics for Precision Medicine (TOPMed) Program (https://www.nhlbiwgs.org/). Use of WGS greatly reduces imputation uncertainty and vastly increases coverage of variation across the human genome. In this study, we utilized state-of-the-art WGS in conjunction with DNA methylation measured by commercial arrays to quantify SNP-CpG associations in over 4,000 FHS participants. Our primary goal was to create a robust mQTL resource to better understand the genetic architecture of DNA methylation and facilitate the discovery of molecular mechanisms underlying a variety of diseases. We also provide examples of how mQTLs can be used in colocalization and Mendelian randomization (MR) analyses to infer the causal roles of DNA methylation in relation to disease phenotypes, with a focus on cardiovascular disease (CVD) risk factors and severity of coronavirus disease 2019 (COVID-19).

## Results

### Participant characteristics

As shown in **Table 1**, our pooled analysis included 4,126 participants (2,320 with DNA methylation data from the 450K array and 1,806 with data from the EPIC array). In the FHS Offspring cohort, blood samples used for the 450K array measurements were collected ∼6 years earlier than those for the EPIC array measurements, while blood samples for both arrays in the Third Generation cohort were obtained at the same visit. Therefore, the mean age for participants with EPIC array data was older than that for the 450K array. There were no substantial differences in sex, BMI, or other CVD risk factors.

**Table 1.** Characteristics of the study population.

|  | Third Generation cohort | Offspring cohort | Omni cohort |
| --- | --- | --- | --- |
| N | 1,945 | 2,129 | 52 |
| Women, % | 52.7 | 54.7 | 46.2 |
| Age, years | 46 ± 8 | 67 ± 9 | 70 ± 9 |
| BMI: kg/m<sup>2</sup> | 27.9 ± 5.7 | 28.2 ± 5.3 | 27.8 ± 4.9 |
| Systolic blood pressure, mm Hg | 116 ± 14 | 129 ± 17 | 126 ± 16 |
| Diastolic blood pressure, mm Hg | 74 ± 9 | 73 ± 10 | 68 ± 10 |
| Hypertension, % | 33.9 | 53.2 | 42.3 |
| Fasting glucose, mg/dL | 96.7 ± 20.8 | 106.3 ± 22.1 | 100.5 ± 12.6 |
| Diabetes, % | 6.0 | 14.7 | 9.6 |
| Triglyceride, mg/dL | 112 ± 79.3 | 119.8 ± 71.9 | 94.9 ± 43.9 |
| High density lipoprotein, mg/dL | 59.3 ± 17.2 | 57.3 ± 18.1 | 65.5 ± 19.3 |
| Low density lipoprotein, mg/dL | 104.5 ± 29.4 | 104.8 ± 31.1 | 87.9 ± 28.9 |

Mean ± SD

### mQTL mapping

In pooled analysis, we identified 94,362,817 *cis*-mQTL variant-CpG pairs for 210,156 unique autosomal CpGs and at *P*<1e-7 and 33,572,145 *trans*-mQTL-CpG pairs for 213,606 unique autosomal CpGs at *P*<1e-14. The numbers of *cis*- and *trans*-mQTL variant-CpG pairs for each chromosome are presented in **Supplemental Table 1**. The *cis*-mQTL variants accounted for 0.7% to 79.9% (median 1.6%) of heritability of DNA methylation, and *trans*-mQTLs accounted for 1.4% to 78.7% (median 2.1%) of heritability. There were 1,080,716 *cis*-mQTL variants, associated with 31,422 unique CpGs (2,345,086 or 2.5% of the 94,362,817 *cis*-mQTL variant-CpG pairs), that accounted for ≥20% of heritability of DNA methylation at the corresponding CpGs (**Figure 2**). We also observed that 185,167 *trans*-mQTL variants accounted for ≥20% of heritability of DNA methylation for 2,711 unique CpGs (314,660 or 0.9% of the 33,572,145 *trans*-mQTL variant-CpG pairs; **Figure 2**). The array-specific results are presented in **Supplemental Table 2**.

We examined whether mQTL variant-CpG pairs identified by other studies^16,17^ were significant in our dataset. For the top independent *cis*-mQTL variant-CpG pairs (168,675 pairs for 104,619 CpGs; *P*<1e-7) identified by the pooled analysis in the five Dutch biobanks,^16^ 66.1% of the pairs (111,557 pairs for 79,099 CpGs) overlapped with our *cis*-mQTL variant-CpG pairs (*P*<1e-7 with consistent effect direction). For the top independent *trans*-pairs (5,865 pairs for 2,066 CpGs with *P*<1e-14) in the Dutch biobanks, 38.4% of the pairs (2,250 pairs for 866 CpGs) overlapped with our *trans*-mQTL variant-CpG pairs (*P*<1e-14 with consistent effect direction). Hawe et al. identified 10,346,172 *cis*- and 819,387 *trans*-mQTL variant-CpG pairs at *P*<1e-14 in a cross-ancestry analysis.^17^ Compared to their study, at *P*<1e-14, we identified 41,224,533 more *cis*-mQTL variant-CpG pairs and 32,752,758 more *trans*-mQTL variant-CpG pairs. Among the 10,346,172 *cis*- and 819,387 *trans*-mQTL variant-CpG pairs reported by Hawe et al.,^17^ 78.3% (n=8,105,456) of *cis*-mQTL variant-CpG pairs and 66% (n=540,851) of *trans*-mQTL variant-CpG pairs were significant and had consistent effect direction in our mQTL database, respectively.

### GO analysis for cis- and trans-mQTLs

Using the top 1,000 unique *cis*-mQTL variants from the pooled analysis, we identified 19 significant GO pathways (16 for Biological Process and 3 for Cellular Component) at FDR<0.05 (**Supplemental Table 3**); the top Biological Process term was dendrite development (GO:0016358; *P*=8.8e-7; FDR=0.01) and the top Cellular Component term was cell periphery (GO:0071944; *P*=3.4e-6; FDR=0.01). The top 1,000 unique trans-mQTL variants from the pooled analysis were linked to nine significant GO pathways (six for Biological Process and three for Cellular Component) at FDR<0.05 (**Supplemental Table 4**); the top Biological Process term was cellular component organization or biogenesis (GO:0071840; *P*=7.8e-7; FDR=0.009) and the top Cellular Component term was cytoplasm (GO: 0005737; *P*=3.4e-6; FDR=0.009).

### Enrichment analysis of mQTL GWAS signals

We examined 9,395,367 *cis*-mQTL variants and 5,820,451 *trans*-mQTL variants located in all autosomal chromosomes identified by the pooled analysis. The enrichment analysis showed that, at FDR<0.05, the *cis*-mQTL variants were enriched with GWAS SNPs associated with 783 traits, representing 27.1% of the traits included in the GWAS Catalog.^7^ For example, we found enrichment of SNPs associated with BMI (enrichment *P*=0 for BMI; **Supplemental Table 5**), systolic BP (enrichment *P*=0), triglyceride level (enrichment *P*=3.5e-231), type 2 diabetes (enrichment *P*=2.4e-194), and coronary artery disease (enrichment *P*=4.8e-118). Compared to the *cis*-mQTL variants, the number of enriched GWAS traits for the *trans*-mQTL variants was lower with enrichment for nine GWAS traits (**Supplemental Table 6**).

### Colocalization analysis

We tested 1,258 CVD risk factor-associated CpGs for colocalization with five CVD-related traits. We found that 104 unique CpGs colocalized with at least one CVD-related traits at PPFC threshold ≥0.7 (overall 155 colocalized pairs; **Supplemental Table 7**). In **Table 2**, we present the top two CpGs that colocalized with each CVD-related trait. For example, cg11554650 (*PPP1R18*), a BMI-associated CpG on chromosome 6, colocalized with type 2 diabetes at SNP rs2516396 (PPFC=0.98), which explained 100% of the observed PPFC; cg05337441 (*APOB*), an LDL-associated CpG at chromosome 2, colocalized with coronary artery disease at rs668948 (PPFC=0.8), which explained 41% of the observed PPFC; and cg03676485 (*LFNG*), a HDL-associated CpG at chromosome 7, colocalized with systolic and diastolic BP at rs4632959 (PPFC=0.99), which explained ∼100% of the observed PPFC.

**Table 2.**
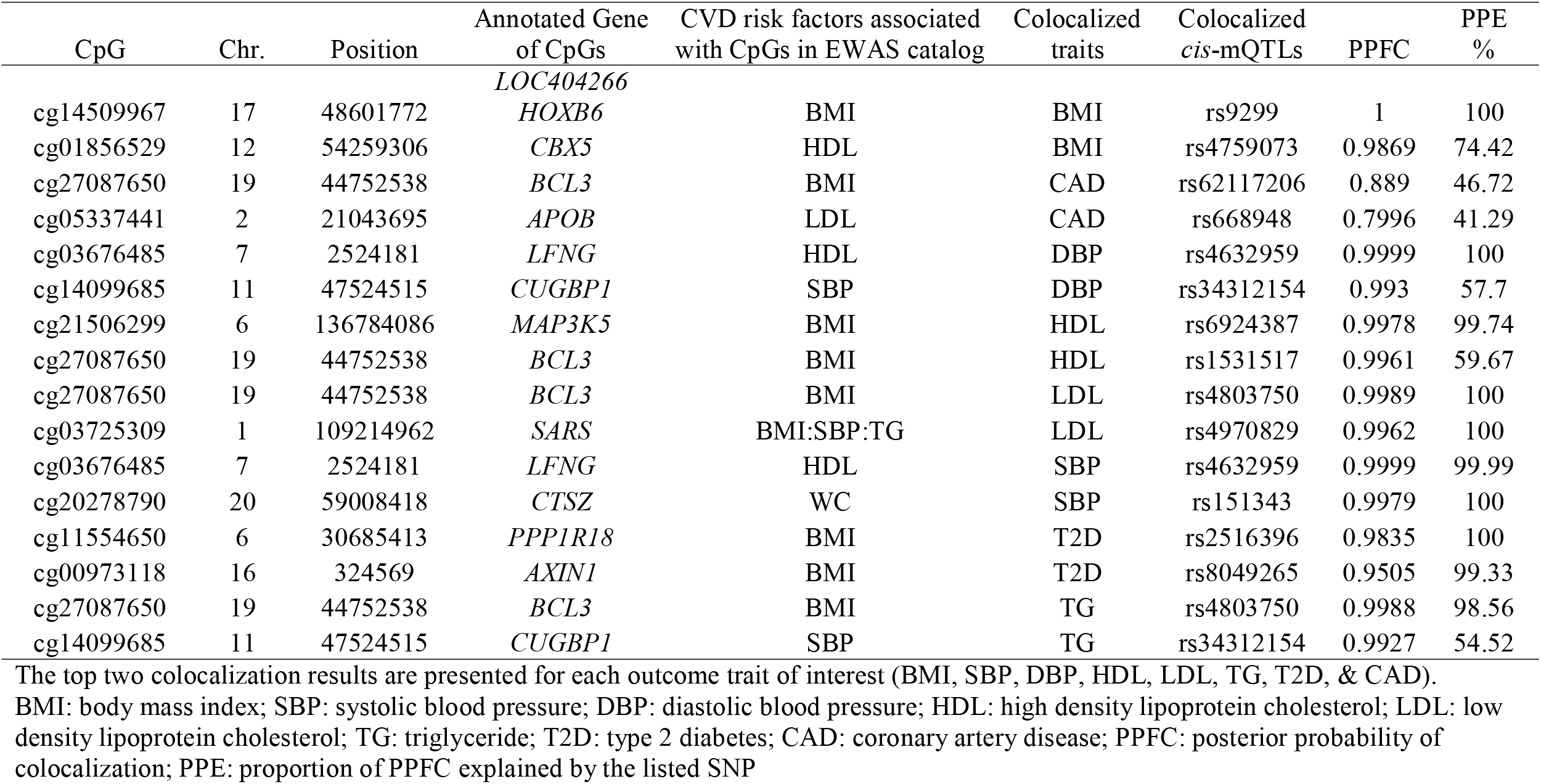
Top colocalization analysis results for CVD risk factor-associated CpGs.

### Mendelian randomization analysis

Using the *cis*-mQTL variants for the 1,258 CVD risk factor-associated CpGs (*P*<1e-6) reported in the EWAS catalog, we conducted MR analysis to test for putatively causal relations of CVD risk factor-associated CpGs with the corresponding CVD risk factors (e.g., HDL-associated-CpGs with HDL and fasting glucose associated-CpGs with type 2 diabetes). After Bonferroni correction for the number of tests in analysis for each trait (e.g., 0.05/566 or 8.8e-5 in analysis for BMI), we identified 58 significant MR associations (**Supplemental Table 8**). The top three CpG-trait pairs were cg05337441 (*APOB*) and LDL (MR effect size: −2.94±0.14, *P*=1.2e-99), cg26663590 (closest gene is *NFATC2IP* in UCSC genome browser) and BMI (MR effect size: −1.39±0.13, *P*=6.3e-26), and cg14099685 (*CUGBP1*) and systolic BP (MR effect size: 138.64±14.85, *P*=9.9e-21). We also demonstrated that 17 CVD risk factor-CpGs were associated with coronary artery disease (**Table 3**; corresponding *P*<3.9e-5), e.g., cg08129017 (SREBF1; reported as associated with BMI and triglyceride in the EWAS catalog; MR effect size: 1.81±0.25, *P*=5e-13) and cg02050917 (*SKI*; BMI-associated CpG; MR effect size: 2.65±0.39, *P*=1.4e-11).

**Table 3.** Mendelian randomization analysis for CVD risk factor-associated CpGs (exposure) in relation to coronary artery disease (outcome)

| CpG | Chr. | Position | Gene | N of IVs | Beta | SE | P |
| --- | --- | --- | --- | --- | --- | --- | --- |
| cg08129017 | 17 | 17825345 | *SREBF1* | 20 | 1.81 | 0.25 | 5.0e-13 |
| cg00184953 | 6 | 31178444 | *PSORS1C3* | 7 | -4.86 | 0.70 | 4.7e-12 |
| cg02050917 | 1 | 2242131 | *SKI* | 13 | 2.65 | 0.39 | 1.4e-11 |
| cg05337441 | 2 | 21043695 | *APOB* | 17 | 1.26 | 0.19 | 3.5e-11 |
| cg21587837 | 6 | 31558116 | *NFKBIL1* | 33 | -2.50 | 0.41 | 1.3e-9 |
| cg03725309 | 1 | 109214962 | *SARS* | 1 | 12.59 | 2.25 | 2.2e-8 |
| cg04545296 | 12 | 48351459 | *ZNF641* | 29 | -0.89 | 0.16 | 2.6e-8 |
| cg26562921 | 16 | 84726822 | *USP10* | 18 | 1.59 | 0.29 | 6.5e-8 |
| cg20544516 | 17 | 17813868 | *MIR33B;SREBF1* | 1 | 7.50 | 1.43 | 1.5e-7 |
| cg21242002 | 4 | 3263352 | *C4orf44* | 4 | -4.93 | 0.96 | 2.7e-7 |
| cg08244301 | 19 | 17499941 | *SLC27A1* | 4 | 3.71 | 0.75 | 7.1e-7 |
| cg21053741 | 6 | 31558083 | *NFKBIL1* | 7 | -3.39 | 0.70 | 1.2e-6 |
| cg27087650 | 19 | 44752538 | *BCL3* | 1 | 7.36 | 1.63 | 6.1e-6 |
| cg10101600 | 2 | 43251603 | *THADA* | 2 | 6.65 | 1.53 | 1.4e-5 |
| cg19224164 | 4 | 2964656 | *GRK4;NOP14* | 13 | -1.67 | 0.40 | 2.3e-5 |
| cg18933331 | 1 | 109643795 |  | 2 | -5.92 | 1.42 | 3.0e-5 |
| cg12467090 | 1 | 204490010 | *PIK3C2B* | 3 | -4.65 | 1.12 | 3.3e-5 |

IV: instrument variables

A recent study conducted in 407 patients with COVID-19 showed that DNA methylation levels at 23 CpGs (located in 20 genes) were associated with COVID-19 severity.^18^ We found that ten of the 23 COVID-19 severity-associated CpGs had at least one *cis*-mQTL variant in our database. We used independent *cis*-mQTL variants (linkage disequilibrium R^2^<0.1), which overlapped with the SNPs tested by the two COVID-19 severity GWAS,^19,20^ to conduct MR analyses. As shown in **Table 4**, we observed that three CpGs, cg14893161 (*PM20D1*; *P*=6e-5 and 0.002 for the two COVID GWAS, respectively), cg17178900 (*PM20D1*; *P*=7e-4 and 0.008), and cg14859874 (*UBAP2L*; P=0.002 and 2e-4), were causally associated with COVID-19 severity after Bonferroni correction in analyses using both COVID GWAS databases.

**Table 4.** Mendelian randomization analysis examining putatively causal relations of COVID-19 severity-associated CpGs to COVID-19 severity.

| CpG | CHR. | BP | Gene | COVID-19 Host Genetics Initiative<br>GWAS (release 6) |  |  |  | GenOMICC study |  |  |  |
| --- | --- | --- | --- | --- | --- | --- | --- | --- | --- | --- | --- |
|  |  |  |  | N of IVs | Beta | SE | P | N of IVs | Beta | SE | P |
| cg07796016 | 1 | 152779584 | *LCE1C* | 19 | 0.08 | 0.26 | 0.76 | 22 | -0.35 | 0.28 | 0.20 |
| cg14859874 | 1 | 154238265 | *UBAP2L* | 36 | 0.64 | 0.21 | 0.002 | 37 | 0.65 | 0.18 | 0.0002 |
| cg17515347 | 1 | 159047163 | *AIM2* | 13 | 1.13 | 0.46 | 0.02 | 13 | 0.34 | 0.44 | 0.44 |
| cg17178900 | 1 | 205818956 | *PM20D1* | 31 | -0.54 | 0.16 | 0.0007 | 31 | -0.46 | 0.17 | 0.008 |
| cg14893161 | 1 | 205819251 | *PM20D1* | 34 | -0.69 | 0.17 | 6.1e-05 | 38 | -0.54 | 0.17 | 0.002 |
| cg08309069 | 6 | 31240651 | *HLA-C* | 46 | -0.40 | 0.19 | 0.04 | 45 | 0.31 | 0.28 | 0.28 |
| cg05030953 | 6 | 31241000 | *HLA-C* | 50 | -0.24 | 0.15 | 0.10 | 43 | 0.47 | 0.18 | 0.01 |
| cg02872426 | 6 | 110736772 | *DDO* | 28 | 0.77 | 0.27 | 0.004 | 27 | 0.39 | 0.28 | 0.16 |
| cg12682382 | 8 | 74787918 | *UBE2W* | 28 | 0.18 | 0.17 | 0.29 | 27 | 0.03 | 0.19 | 0.88 |
| cg13571460 | 9 | 124989337 | *LHX6* | 17 | 0.10 | 0.35 | 0.78 | 18 | -0.21 | 0.33 | 0.53 |

IV: instrument variables

## Discussion

To create a cutting-edge genome wide resource of *cis-* and *trans*-mQTLs, we analyzed whole genome sequences in conjunction with array-based DNA methylation data from 4,126 FHS participants. Our pooled analysis identified 94,362,817 *cis*-mQTL variant-CpG pairs (9,395,367 *cis*-mQTL variants; 210,156 unique autosomal CpGs; *P*<1e-7) and 33,572,145 *trans*-mQTL variant-CpG pairs (6,039,960 *trans*-mQTL variants; 213,606 unique autosomal CpGs; *P*<1e-14). We found enrichment of mQTL variants for disease-associated SNPs from GWAS. Using *cis*-mQTL variants, colocalization analyses support connections between CpGs with CVD traits. MR analyses further demonstrated that *cis*-mQTLs can be used to test causal relations of CpGs to multiple phenotypes such as CVD traits and COVID-19 severity. A comprehensive summary data set will be posted to the National Heart, Lung, and Blood Institute’s BioData Catalyst site and will be freely accessible to the scientific community. Taken together, our study created a robust mQTL repository to better understand the epigenetic mechanisms underlying a wide range of diseases.

Consistent with our previous mQTL study^15^ and others,^21,22^ a majority of SNP-CpG pairs are *cis*. For example, the number of *cis*-mQTL-CpG pairs was 2.8 times of that of *trans*-mQTL-CpG pairs in our pooled analysis (1.5 times using *P*<1e-14). To the best of our knowledge, our study is the largest mQTL mapping project using WGS, including ∼20 million SNPs and INDELs and ∼850 thousand CpGs. Our database expands the existing literature by adding ∼40 million novel *cis*- and ∼30 million *trans*-mQTL-CpG pairs based on WGS rather than imputed genotypes from array-based genotyping. In addition, our database included *cis*- and *trans*-mQTLs for 180,692 unique CpGs present on the EPIC array that are not on the 450K array. Compared to the older 450K array, the EPIC array increases CpG coverage of specific genomic regions such as enhancers and non-coding regions.^23^ Therefore, our data therefore will facilitate future studies that examine the potential biological function and clinical impact of DNA methylation at these genomic regions.

To showcase the application of our mQTL database, we demonstrated the enrichment of mQTL variants for disease-associated SNPs from GWAS using the GWAS Catalog.^7^ For example, analysis utilizing *cis*-mQTL variants showed enrichment for SNPs associated with CVD and multiple CVD risk factors including BMI, systolic BP, triglyceride, type 2 diabetes, and coronary artery disease. Our colocalization analysis using *cis*-mQTL variants for CpGs and GWAS summary statistics of these variants for CAD identified colocalization of an LDL-associated CpG, cg05337441 (*APOB*), with coronary artery disease. A intergenic SNP rs668948, mapped to *APOB* and *TDRD15*, explained 41% of the observed colocalization. The product encoded by *APOB* is the main apolipoprotein of LDL that serves as the ligand for the LDL receptor. The atherogenic potential of apolipoprotein B-100 has been demonstrated by many studies including MR analysis.^24-27^ Our data are consistent with the notion that DNA methylation contributes to the atherogenicity of LDL and suggest that future studies are needed to examine the exact molecular underpinnings of these observations. Also, in line with these observations, our MR analysis showed that many CVD risk factor-associated CpGs are putatively causal for CVD and CVD risk factors (Supplemental Table 8). These findings provide epigenetic insights into associations reported in GWAS. For example, we observed that cg12816198 (*IRF5*) was associated with systolic BP (MR *P*=6.3e-8). SNP rs4728142, an intergenic variant mapped to genes *IRF5* and *KCP*, has been reported to be associated with hypertension in previous GWAS.^28^ This SNP (rs4728142) is a strong *cis*-mQTL variant for cg12816198 (*IRF5*; *P*=7e-215) and the leading instrumental variable in the MR analysis for systolic BP (single SNP MR analysis *P*=2.7e-9), suggesting a causal pathway whereby rs4728142 modifies DNA methylation levels at cg12816198 with downstream effects on systolic BP. Interestingly, both colocalization analysis and MR analysis showed a connection between cg27087650 (*BCL3*) and coronary artery disease through *cis*-mQTL variant rs62117206 (intronic to *BCL3*; *P*=3.6e-15; linkage disequilibrium R^2^ =1 with rs4803750, another *cis*-mQTL variant of cg27087650; *P*=1.8e-14). CpG cg27087650 is located in the gene body of *BCL3*, which encodes a protein functioning as a transcriptional co-activator through its association with NF-kappa B homodimers. Expression of *BCL3* has been linked to CVD and cancer.^29-31^These examples provide proof of principle that integrating *cis*-mQTLs with CpGs and traits can reveal biological pathways by linking DNA methylation to a variety of diseases.

Our mQTL database can also be used to screen candidate DNA methylation sites for further consideration in experimental and interventional studies. This is exemplified by our MR analysis that revealed a putatively causal effect of COVID-19 associated CpGs on disease severity. Our COVID analysis focused on ten CpGs that were identified in a case-control study of COVD-19 severity.^18^ Because of the retrospective design of the study,^18^ it could not infer causal relations between DNA methylation at these CpGs and the severity of COVID-19. Our analysis highlighted three COVID-related CpGs annotated to genes *PM20D1* and *UBAP2L* that were putatively causal for COVID-19 severity; more research is needed to understand if and how these CpGs might influence outcome in patients with the COVID-19.

In parallel with our mQTL project, our research team is examining eQTLs and expression quantitative trait methylation sites (eQTM) using WGS, RNA sequencing, and DNA methylation resources obtained in FHS participants. The eQTL and eQTM resources are also freely available online via the BioData Catalyst site. These molecular resources enable users to explore how DNA methylation affects transcriptional activities and pathways leading to a wide range of disease phenotypes. These molecular resources can be used in concert to reduce bias due to reverse causality and unmeasured confounding, particularly environmental confounders.^32,33^ Nonetheless, this study has several limitations that warrant discussion. Our analysis was conducted in a group of middle-aged and older, primarily white adults; therefore, the findings in this study may not be generalizable to other populations. Nonetheless, we demonstrated that mQTLs identified in other studies,^16,17^ including those identified in a cross-ancestry analysis,^17^ were well replicated in our database. We captured whole blood-based DNA methylation profiles, which can serve as candidate biomarkers for diseases; however, they may not reflect tissue-specific DNA methylation levels, which may be relevant to specific diseases. MR analysis was used to showcase the potential application of our mQTL database; however, MR analysis is based on assumptions that may not be testable.^34^ Also, DNA methylation can be affected by both genetic and environmental factors. We did not attempt to test effect modification by environmental factors in this study. Future studies with larger sample sizes in diverse population are needed to replicate and expand our mQTL resource.

In conclusion, we have identified millions of *cis*- and *trans*-mQTL variant CpG pairs using state-of-the-art WGS data in conjunction with high-throughput DNA methylation data. We demonstrated the utility of this vast mQTL resource by conducting GWAS signal enrichment analyses, colocalization, and MR analyses. Our mQTL repository is freely available via the NCBI Molecular QTL Browser for the scientific community to study the role of DNA methylation in health and disease.

## Methods

### Study Population

The study sample included consenting participants from the FHS Offspring, Third Generation, and Omni cohorts. In 1971, the FHS recruited the offspring of participants in the Original FHS cohort as well as the spouses of offspring to form the FHS Offspring cohort.^35^ The children of the Offspring cohort participants were recruited to the Third Generation cohort beginning in 2002.^36^ Omni cohorts were established in parallel with the Offspring and the Third Generation cohorts. In the current investigation, the study sample included 4,126 FHS participants with whole blood derived DNA methylation and WGS data; 2,129 participants in the Offspring cohort (exam 8, N=869; exam 9, N=1,260), 1,945 participants in the Third Generation cohort (exam 2), and 52 participants in the Omni cohort. The FHS protocols and procedures were approved by the Institutional Review Board for Human Research at Boston University Medical Center, and all participants provided written informed consent. All research was performed in accordance with relevant guidelines/regulations.

### Study design

A flow chart of the study design is presented in **Figure 1**. The FHS had two sets of DNA methylation data, one set included 3,460 participants assayed with the Illumina BeadChip 450K (450K array; 2,009 Offspring exam 8 participants and 1,451 Third Generation exam 2 participants) and the second set included 1,806 participants assayed with the Illumina EPIC array (EPIC array; 1,260 Offspring exam 9 participants, 494 Third Generation exam 2 participants, and 52 Omni cohort participants). To maximize the sample size, as our primary analysis we conducted a pooled analysis of the two data sets. Of note, 1,140 Offspring participants were included in both sets, i.e., these participants had 450K array-based methylation data from exam 8 and EPIC array-based methylation data from exam 9. In the pooled analysis, we selected the EPIC array-based data for these 1,140 participants to avoid any duplication. We also conducted array-specific analysis to explore if mQTLs were replicable and to examine mQTLs that are unique to the EPIC array. We then examined the top *cis*- and *trans*-mQTLs by conducting Gene Ontology (GO) pathway analysis and enrichment analysis. We tested *cis*-mQTLs for colocalization and causal association using two-sample MR analysis with CVD traits and COVID-19 severity.

**Figure 1.**
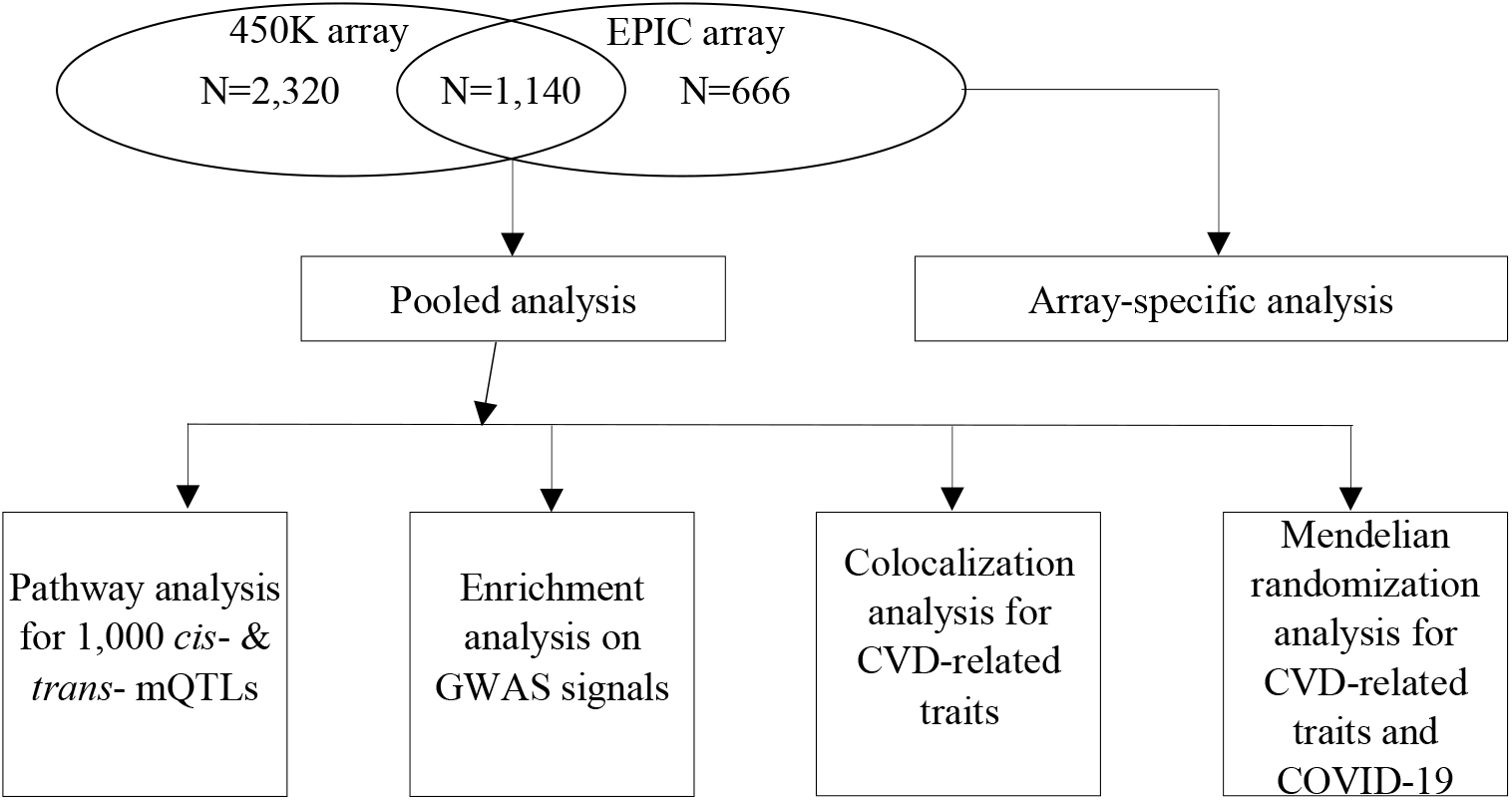
Study design.

**Figure 2.**
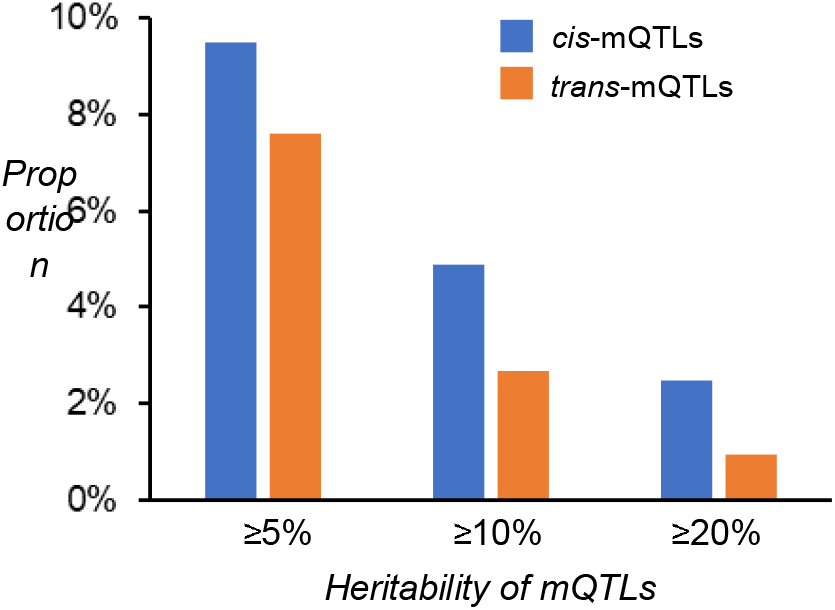
Heritability of DNA methylation explained by the *cis*- and *trans*-mQTLs identified in the pooled analysis. The total number of cis-mQTLs is 94,362,817 and the total number of trans-mQTLs is 32,434,987.

### DNA methylation profiling

Preparation of whole blood samples for DNA methylation profiling was conducted as detailed previously.^15^ Briefly, DNA was obtained from whole blood buffy coat samples and prepared using bisulfite conversion before whole-genome amplification, fragmentation, array hybridization, and single-base pair extension. DNA methylation was then measured in 4,170 FHS participants using the Illumina Infinium Human Methylation-450 Beadchip (450K array) in three batches (Batch 1, N=499; Batch 2, N=2,149; and Batch 3, N=1,522). Of these, 3,460 participants also had WGS data. Additionally, the Illumina MethylationEPIC 850K BeadChip (EPIC array) was used in 1,806 FHS participants with WGS. All participants were with missing methylation levels of no more than 5% of CpGs (detection *P*<0.01) and none of them were outliers in a multi-dimensional scaling plot. The CpGs have been prefiltered so that all CpGs had < 5% missing values (detection *P*<0.01). We calculated DNA methylation beta values (range 0 to 1) as the ratio of mean methylated and sum of methylated and unmethylated probe signal intensities. We used the DASEN method^37,38^ to normalize the methylation beta values.

### Whole Genome Sequencing

WGS of FHS participants was performed by the Broad Institute as part of the NHLBI’s TOPMed program.^39^ Genomic DNA from whole blood samples from 2,194 FHS Offspring cohort and 1,582 Third Generation cohort participants was sequenced at >×30 depth of coverage.^39^ Genetic variations were identified in a joint calling of all samples by the TOPMed Informatics Resource Center at University of Michigan. Centralized read mapping, genotype calling, and quality control were also performed at the TOPMed Informatics Research Center. This analysis used genetic variants generated from TOPMed Freeze 10a. We analyzed 20,696,115 SNPs and insertion/deletion polymorphisms (INDELs) with minor allele count (MAC) ≥10. WGS data acquisition is described on the Database of Genotype and Phenotype (dbGaP) website (https://www.ncbi.nlm.nih.gov/projects/gap/cgi-bin/study.cgi?study_id=phs000974.v4.p3).

### mQTL mapping

The mQTL mapping was conducted separately for DNA methylation data generated using the 450K array and the EPIC array. In the analysis for the 450K array data, we calculated residuals for methylation beta values obtained within each of the three methylation batches after adjusting for relevant technical covariates. Whereas, in the analysis for the EPIC array data, we derived residuals using all available samples, also adjusting for technical covariates. The residuals from separate datasets were then combined. We then used linear regression models to perform the association analyses between the SNPs and the CpGs, adjusting for sex, age, differential leukocyte counts (estimated using the Houseman method^40^), along with the top 15 residual methylation principal components (PCs) and five genetic PCs. We chose to adjust for 15 methylation PCs and five genetic PCs because this strategy resulted in the highest replication rate between the 450K array data and the EPIC array data. Because of relatedness among FHS study participants, linear mixed models were used in mQTL mapping to account for family structure. The primary pooled analysis examined 452,567 CpGs that are common to both arrays. The 450K array-specific analysis analyzed the same 452,567 CpGs and the EPIC array-specific analysis examined 413,524 additional CpGs (i.e., CpGs not included in the pooled analysis). For *cis*-mQTLs (defined as within 1 million base pairs from the CpG site), we considered two-sided *P*<1e-7 to be statistically significant, whereas for *trans*-mQTLs (defined as located ≥1 million base pairs away from the CpG site or on a different chromosome), we considered two-sided *P*<1e-14 to be statistically significant. The *P* value thresholds were selected for *cis*-mQTLs based on the Bonferroni correction for the number of CpGs tested (i.e., n=452,567) and for *trans*-mQTLs based on internal discovery-validation experiment that gave the highest *trans* replication rate.

### mQTL Replication

To explore consistency between our mQTLs with published databases, we examined mQTLs identified in two large studies, one conducted by Bonder et al in 3,841 individuals from five Dutch biobanks^16^ and the other conducted by Hawe et al in 3,799 European individuals and 3,195 individuals from South Asia.^17^ Both studies analyzed SNPs based on commercial arrays with imputation. Because the number of SNPs analyzed in the two studies (∼5 and ∼9 million, respectively) was smaller than that tested in the present study (∼20 million), we examined whether mQTLs identified in the two studies were also significant in our database.

### Gene Ontology analysis

We tested the over-representation of GO terms based on genes annotated to the top 1,000 *cis*-mQTL variants (for 1,000 CpGs) with Entrez IDs identified by the pooled analysis. The default setting in the *goana* function from the R *limma* (Linear Models for Microarray and RNA-seq Data) package was used to conduct the GO analysis.^41^ GO terms (Biological Process, Cellular Component, and Molecular Function) with false positive rate (FDR) <0.05 were reported. We repeated the same analysis for the top 1,000 *trans*-mQTL variants.

### GWAS Enrichment analysis

We analyzed all SNPs with association *P*<5e-8 included in the NHGRI-EBI GWAS Catalog (https://www.ebi.ac.uk/gwas/).^7^ We identified 243,587 entries for 2,960 GWAS traits. In this analysis, we examined all mQTL variants with unique RSIDs in *cis* or *trans* at *P*<1e-7 or *P*<1e-14, respectively. Fisher’s exact test was used to perform the enrichment analysis for each trait, and traits with FDR<0.05 were reported.

### Colocalization analysis

We conducted colocalization analysis using the R *HyPrColoc* package, a highly efficient deterministic Bayesian algorithm based on GWAS summary statistics.^42^ We reported the posterior probability of full colocalization (PPFC). Default prior configuration parameters (*prior*.*1*=1e-4 and *prior*.*c*=0.02) and threshold of 0.7 for PPFC were used. We extracted *cis*-mQTL variants (*P*<1e-7) derived from the present pooled analysis for 1,258 CpGs associated with CVD risk factors in the EWAS catalog (*P*<1e-6) including BMI, waist circumference, fasting glucose, systolic blood pressure (systolic BP), diastolic blood pressure (diastolic BP), high-density lipoprotein cholesterol (HDL), low-density lipoprotein cholesterol (LDL), and triglyceride.^6^ We examined the colocalization of these CVD risk factor-associated CpGs with CVD-related traits including BMI, BP, lipid concentrations, type 2 diabetes, and coronary artery disease. Summary statistics for associations between *cis*-mQTL variants and GWAS SNPS for CVD-related traits were obtained from published GWAS databases.^27,43-47^

### Mendelian randomization analysis

To showcase the potential use of the mQTL resource in causal inference analyses, we conducted MR analyses to infer causal associations of the CpGs with the abovementioned CVD-related traits and COVID-19 severity. In the MR analysis for CVD-related traits, we used the same *cis*-mQTL variants for the 1,258 CVD risk factors. COVID-19-associated CpGs were obtained from a recently published EWAS of COVID-19 severity.^18^ We performed MR analyses using a two-sample MR approach.^48^ We used independent *cis*-mQTL variants with pair-wise linkage disequilibrium (LD) r^2^ <0.1 as instrumental variables (IVs). Using the *TwoSampleMR* R package,^49^ we performed the primary analysis using the inverse variance weighted (IVW) method and sensitivity analysis using the MR-Egger method. We tested for potential horizontal pleiotropy by examining the MR-Egger intercept *P* value. The effect sizes and standard errors for IV-CpG associations were obtained from the pooled mQTL analysis. The effect sizes and standard errors for associations between IVs and CVD-related traits were obtained from the published large GWAS described above.^27,43,44,46,47^ We obtained effect sizes and standard errors from two GWAS for COVID-19 severity conducted by the COVID-19 Host Genetics Initiative^20^ and the Genetics of Mortality in Critical Care (GenOMICC) study.^19^ The COVID-19 Host Genetics Initiative included 8,779 cases (death or hospitalization requiring respiratory support due to COVID-19) and 1,001,875 population controls and the GenOMICC study included 7,491 cases (confirmed COVID-19 requiring continuous cardiorespiratory monitoring in intensive care units) and 48,400 population controls.

## Data Availability

All data produced are available online at dbGap

https://www.ncbi.nlm.nih.gov/projects/gap/cgi-bin/study.cgi?study_id=phs000974.v4.p3

## Notes

**Funding:** The Framingham Heart Study was supported by NIH contracts N01-HC-25195, HHSN268201500001I, and 75N92019D00031. DNA methylation assays were supported in part by the Division of Intramural Research (D. Levy, Principal Investigator) and an NIH Director’s Challenge Award (D. Levy, Principal Investigator). The analytical component of this project was funded by the NHLBI Division of Intramural Research (D. Levy, Principal Investigator). Whole genome sequencing for the TransOmics in Precision Medicine (TOPMed) program was supported by the NHLBI. Core support including centralized genomic read mapping and genotype calling, along with variant quality metrics and filtering were provided by the TOPMed Informatics Research Center (3R01HL-117626-02S1; contract HHSN268201800002I). Core support including phenotype harmonization, data management, sample identity QC, and general program coordination were provided by the TOPMed Data Coordinating Center (R01HL-120393; U01HL-120393; contract HHSN268201800001I). J. Ma is supported by NIH grants, K22HL135075 and R01AA028263. The authors’ contributions were as follows— DL and JM designed research, interpreted results, and had primary responsibility for final content; RJ and JM conducted the analyses; JM and RJ wrote the manuscript; HB contributed to the literature review used in the paper; CL AK, HS, HB, BT, MS, PJM, DYC, PC, NLH, ANP, MF, NS, RSV, and TH critically reviewed the manuscript; and all authors read and approved the final manuscript. The authors declare no conflicts of interest.

### Competing Interest Statement

The authors have declared no competing interest.

### Funding Statement

The Framingham Heart Study was supported by NIH contracts N01-HC-25195, HHSN268201500001I, and 75N92019D00031.

### Author Declarations

Institutional Review Board for Human Research at Boston University Medical Center

### Summary of Updates

Summary data storage site has been updated.

